# Phenotypic signatures in clinical data enable systematic identification of patients for genetic testing

**DOI:** 10.1101/2020.07.21.20159491

**Authors:** Theodore J. Morley, Lide Han, Jonathan Morra, Nancy J. Cox, Lisa Bastarache, Douglas M. Ruderfer

**Affiliations:** Division of Genetic Medicine, Department of Medicine, Vanderbilt University Medical Center, Nashville, TN, USA; Vanderbilt Genetics Institute, Vanderbilt University Medical Center, Nashville, TN, USA; Zefr, Los Angeles, CA, USA; Center for Precision Medicine, Department of Biomedical Informatics, Vanderbilt University Medical Center, Nashville, TN, USA; Department of Psychiatry and Behavioral Sciences, Vanderbilt University Medical Center, Nashville, TN, USA

## Abstract

Around five percent of the population is affected by a rare disease, most often due to genetic variation. A genetic test is the quickest path to a diagnosis, yet most suffer through years of diagnostic odyssey before getting a test, if they receive one at all. Identifying patients that are likely to have a genetic disease and therefore need genetic testing is paramount to improving diagnosis and treatment. While there are thousands of previously described genetic diseases with specific phenotypic presentations, a common feature among them is the presence of multiple rare phenotypes which often span organ systems. Here, we hypothesize that these patients can be identified from longitudinal clinical data in the electronic health record (EHR). We used diagnostic information from the EHRs of 2,286 patients that received a chromosomal microarray and 9,144 matched controls to train and test a prediction model. We identified high prediction accuracy (AUROC = 0.97, AUPR = 0.92) in a held-out test sample and in 172,265 hospital patients where cases were defined broadly as interacting with a genetics provider (AUROC = 0.9, AUPR = 0.63). High probabilities (median = 0.97) were associated with 46 patients carrying a known pathogenic copy number variant (CNV) among a subset of 6,445 genotyped patients. Our model identified many more patients needing a genetic test while increasing the proportion having a putative genetic disease compared to the current nonsytematic approach. Taken together, we demonstrate that phenotypic patterns representative of a genetic disease can be captured from EHR data and provide an opportunity to systematize decision making on genetic testing to speed up diagnosis, improve care, and reduce costs.

## Introduction

Rare diseases, of which the majority are genetic, were recently estimated to affect 3.5-6.2% of the world’s population^1,2^. Many genetic diseases have yet to be discovered or characterized, leaving those patients with particularly long, challenging diagnostic odysseys^2,3^. Even for the thousands that have already been described^4,5^, heterogenous clinical symptoms may complicate identification of the underlying cause, delaying a diagnosis and an opportunity for potential medical benefits. Genetic testing represents a standard means to diagnose a patient with a genetic disease. However, current approaches that determine which patients receive a genetic test are inconsistent and inequitable^6^. For numerous conditions where genetic testing is recommended, the vast majority of patients still do not receive a genetic test^7,8^. Developing a systemized way to identify patients likely to have a rare genetic disease could guide genetic testing decision-making to improve diagnostic outcomes, reduce healthcare costs and burden on patients, and enable opportunities for improved care.

The identification of genetic diseases has typically been through clinical ascertainment on shared syndromic features^9,10^. However, there exists variable expressivity and penetrance such that two patients with the same underlying genetic variant may not present similarly or with all or many of the features of the well characterized genetic disease^11^. For example, a large deletion on chromosome 22 causes 22q11.2 deletion syndrome, which includes both velocardiofacial syndrome and DiGeorge syndrome, historically believed to be different syndromes due to differing clinical presentations. Additionally, patients may carry multiple contributing genetic factors leading to a phenotypic presentation that deviates from those previously defined and challenging a clear diagnosis^12,13^.

Longitudinal clinical data stored in the electronic health record (EHR) have enabled approaches to identify patients at risk for numerous conditions^14^. In particular, recent work has shown that specific genetic diseases can be identified by looking for patients carrying many of the expected symptoms^15,16^. While each genetic disease may present with a recognizable phenotypic profile, across the majority of genetic diseases there exists a recurring pattern of multiple phenotypes that are often rare and affect multiple organ systems. We hypothesize that this constellation of rare and diverse phenotypes is a hallmark signature of patients with a genetic disease and can be captured from data in the EHR.

Here, we test this hypothesis by building a machine-learning based prediction model to identify patients that have a clinical profile representative of getting a genetic test for suspicion of having a genetic disease. Specifically, we trained and tested our model on 2,286 patients that received a chromosomal microarray and 9,144 demographically matched controls using only diagnostic information from the EHR. We show highly accurate performance in our held-out testing sample as well as an independent set of over 170,000 hospital patients. We further validate this model’s ability to identify patients with genetic diseases in patients having putative pathogenic copy number variants and those carrying a diverse array of validated genetic diseases including many not present in our training data. Overall, our approach establishes the potential to capture genetic disease patients from EHR data and presents a systemized way to improve the consistency and equity of genetic testing.

## Methods

### Identification of patients receiving a genetic test and matching controls from EHR data

Our case population included 2,388 patients who received a chromosomal microarray (CMA) intended to identify large deletions and duplications. Those receiving this test were identified by CMA pathology reports from 2012 – 2018 from the Vanderbilt University Medical Center (VUMC) Synthetic Derivative (a de-identified EHR system). The extracted data for the CMA reports includes the date of report, indication for receiving the test, and interpretation (whether there were reported variants and if so, the size and location of the variant). Twenty-four percent of patients (575/2,388) had at least one abnormal finding of which the majority (84%) were a gain or loss with the rest being runs of homozygosity or more complex genetic variation. For every case, we identified four patients having identical age, sex, race, number of unique years in which the patient had visited VUMC, and the closest EHR record length in days (maximum of 100 days difference). After matching, there were 2,286 cases and 9,144 controls (Table 1). The vast majority (95%) of the cases were less than 20 years (mean age: 8.1), most were male (61.3%) and white (75.6%).

**Table 1.**
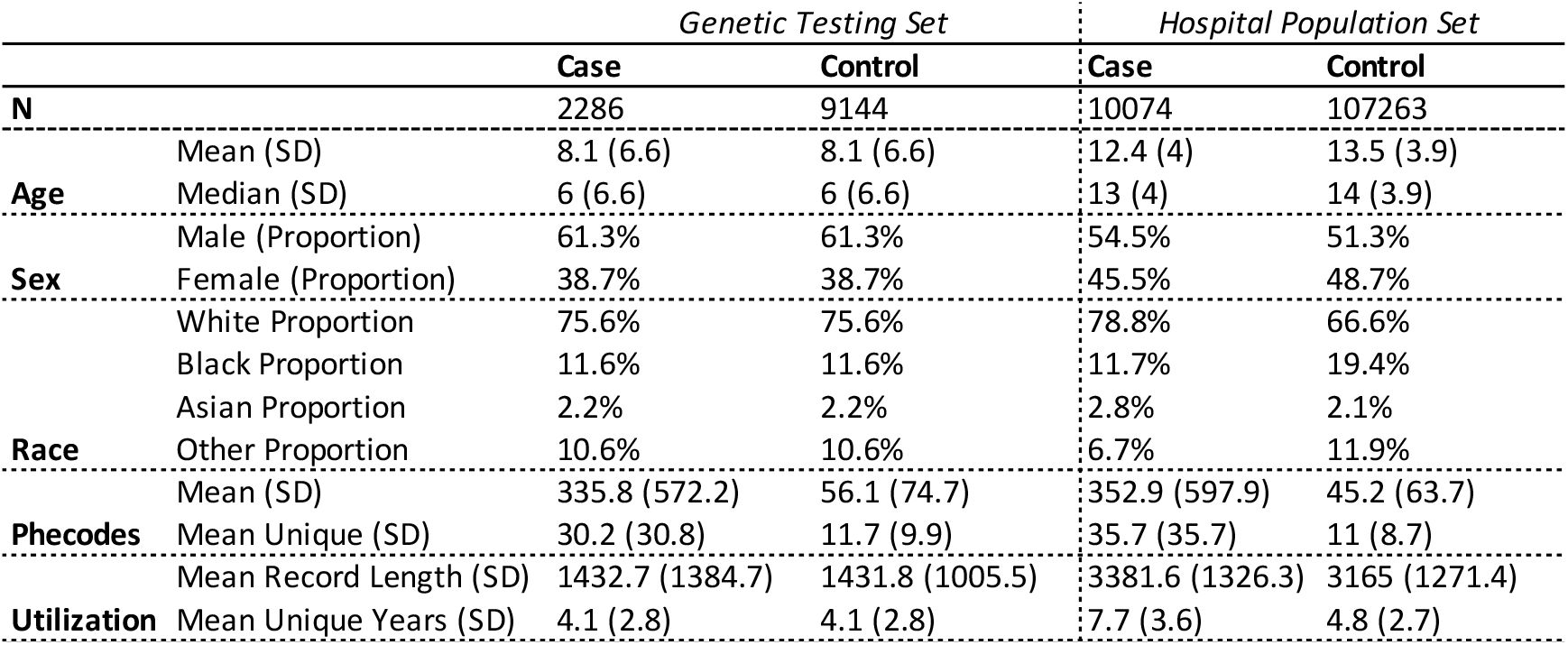
Demographic and hospital utilization information for genetic testing and hospital population datasets.

### Generating features matrices to include in prediction model

We translated ICD9-CM and ICD10-CM codes to 1,685 pheWAS^17^ codes (phecodes, version 1.2) and generated three different methods of representing these patients’ diagnostic data. The first was a binary matrix indicating presence or absence of phecodes, the second was a matrix of phecode counts, and the third was a broadly defined phenotypic risk score (pheRS)^16^. Instead of being disorder specific, we calculated a pheRS across all phecodes, creating a singular score which aims to balance both the diversity of a patient’s phenotypes as well as the rarity of those phenotypes. In calculating prevalence as weights, we rolled all phecodes up the hierarchy to ensure higher level codes were at least as common as the codes below them. For prediction, we removed all phecodes under the category of congenital anomalies, as these codes could be used to indicate a genetic test or a diagnosis from one.

### Building and testing of prediction model

We trained our model using four-fold cross validation on 80% of the data and reserved 20% as a held-out test set. For the binary phecode and phecode count matrices, we additionally evaluated three different methods of dimensionality reduction. They consisted of principal component analysis (PCA), uniform manifold approximation and projection (UMAP)^18^, and PCA preserving a number of components which account for at least 95% of the cumulative variance in the dataset fed into UMAP for final dimensionality reduction. We considered four different classification algorithms on this dataset; naïve Bayes, logistic regression, gradient boosting trees, and random forest. Aside from UMAP, all classification algorithms used were from the scikit-learn package^25^. After selecting a range of hyperparameters for each classifier and dimensionality reduction method we applied a grid search within our cross-validation framework and optimized our model selection on the area under the precision recall curve (average precision) which summarizes all available precision (positive predictive value) for every possible recall (sensitivity).

To assess whether phecodes occurring at or after the time of genetic testing affected performance, we also trained a model censoring from the date of the CMA report onwards. Therefore, the training and testing procedure described above was performed twice. To test potential disparities within our model within race and sex, we trained classifiers through the same process as the main classifier was trained, except that we used only phecode counts matrix as input as it was what performed best in the primary task. We used the same sample set, but the classification target was instead set to sex or race.

### Defining a hospital population dataset for independent validation

We extracted 845,423 VUMC patients with a record length of at least four years. We reduced this sample to 172,265 that were under 20 years of age to best match our training sample. Cases (n=10,074) were defined as those identified as having evidence of being seen in a genetic clinic by searching for relevant keywords such as “genetic” within the titles of their clinical notes or the first 200 characters of the note, excluding notes with titles containing the phrase ‘hereditary cancer’, as this indicated that the note originated from the hereditary cancer genetics clinic. We further performed a broad search for any clinical suspicion of genetic disease in patients’ clinical records to identify patients that may have received genetic tests but who didn’t visit a genetic clinic at VUMC. These patients were identified using regular expressions related to “genet”, “chromosom”, “congenital”, “copy number”, “gene test”, “genetic test”, “nucleotide”, “dna”, “mutation”, “genotype”, “heterozy”, “homozy”, “recessive”, “autosomal dominant”, “exon”, “genes”, and excluding common negations such as “no genet”, “no congenital”, or “not due to genet”. In total, there were 64,924 patients in this category including 99.2% of the cases (n=9,996). After removing those patients, we were left with 107,263 controls to compare to our cases to further validate our model’s performance.

### Copy number variant (CNV) quality control

We used a set of 93,626 patients from the Vanderbilt Biobank that were genotyped on the Illumina Multi-Ethnic global Array (MEGAex) for these studies. To improve quality of input to CNV calling, we reduced the set of total variants (n = 2,038,233 SNPs) to only those with high genotyping call rates (>95%). CNVs were called using PennCNV^19^ with population frequency of B allele (PFB) file and GC model file^20^ generated from 1,200 randomly selected samples. We removed samples where log R ratio standard deviation (LRR SD) <0.3, B allele frequency drift <0.01, and the absolute value of waviness factor (|WF|) < 0.05. Only CNVs greater than 10 kb and having at least 10 contributing variants were retained. We further removed samples with outlier (z-scores greater or less than 1.96) numbers of CNVs after quantile normalization. CNVs were removed if they overlapped genomic regions such as centromeres, telomeres and ENCODE blacklist regions^21^. Adjacent CNVs were merged if gap was less than 20% of the combined length of the merged CNV. Finally, only CNVs in less than 1% of the sample (allele frequency 0.5%) were kept for analysis. There were 945,196 CNVs among 86,294 samples of which 6,445 were among the 172,265 patients in the hospital reference population described above.

### Validation of prediction model using CNVs

Further validation of our model was performed by comparing the CNVs to three sets of pathogenic variants. First, we used a list of 66 pathogenic CNV syndromes from the DECIPHER consortium^22^. We examined individuals who were in our hospital population set and had at least 50% overlap with a CNV classified with grade one pathogenicity. Second, we downloaded 7,773 putative pathogenic CNVs from ClinGen (downloaded from UCSC Genome Browser June 2019) and again required 50% overlap. Finally, we identified 132 patients carrying a 10Mb or greater duplication on chromosome 21 indicative of Down Syndrome.

### Gold standard genetic diagnoses extraction and validation

We used a previously developed cohort of patients with confirmed clinical diagnoses for with 16 different genetic diseases (achondroplasia, alpha-1 antitrypsin deficiency, cystic fibrosis, DiGeorge syndrome, Down syndrome, fragile X syndrome, hemochromatosis, Marfan syndrome, Duchenne muscular dystrophy, neurofibromatosis type I, neurofibromatosis type II, phenylketonuria, polycythemia vera, sickle cell anemia, telangiectasia type I, tuberous sclerosis).^15^ These patients were identified through manual chart review. Using this gold standard cohort of patients diagnosed with genetic disease, we validated the performance of our model by comparing the proportion of patients with the genetic diagnoses and probability above different thresholds to the proportion of the population with probabilities above the same thresholds. In this way we aim to quantify the fold-increase in genetic disease patients that would be identified at particular thresholds compared to the proportion of patients that would be tested.

## Results

### Demographic and phenotype description of genetic testing sample

Our primary case population consisted of 2,286 patients who received a chromosomal microarray (CMA). We matched each CMA patient to 4 controls based exactly on age, sex, race, number of unique years in which they visited VUMC, and the closest available match on medical record length in days (maximum difference of 100 days). The vast majority (95%) of the CMA recipients were less than 20 years old (mean age: 8.1), most were male (61.3%) and white (75.6%, Table 1). Twenty-four percent (n = 550) of patients had an abnormal result reported including 250 with at least one gain, 257 with at least one loss. Among these, 37% (201/550) included a potential diagnosis in the report. While the reported genomic coordinates were most often unique there were several known recurrent syndromes seen more frequently including DiGeorge syndrome, Charcot-Marie Tooth syndrome and 16p11.2 Deletion syndrome. For the 76% of patients where reports were considered “normal” it is important to note that only a small subset of genetic variation was being tested and there is substantial opportunity for other genetic variation to be contributing to the presented symptoms.

We tested the frequency of phecodes^17^ between the CMA patients and the matched controls. Conditions of early development such as autism, developmental delay, delayed milestones, and multiple congenital anomalies such as heart defects represented the most significantly associated phecodes (Supplementary Fiigure 1). When performing the same analysis between CMA patients with an abnormal report vs those without we identified 2 significant phecodes after correction for 1,620 tests (p < 3.1×10^−5^) including chromosomal anomalies (758.1, p = 3.31×10^−151^) and developmental delays and disorders (315, p = 2.73×10^−5^).

### Building a prediction model for patients receiving a CMA

We posed a prediction problem in which we sought to distinguish individuals who received a CMA from matched controls, capturing the clinical suspicion of a genetic disease but in an automated and systemized way. We included both presence/absence of phecodes and counts as input and applied multiple prediction methods including naïve Bayes, logistic regression, gradient boosting trees, and random forest (see Methods). Chromosomal anomalies and all 56 phecodes in the congenital anomalies group were removed to avoid potential bias if those phecodes resulted from the CMA. We further employed several approaches in order to reduce dimensionality of our input and included an all phecode phenotype risk score^16^ for comparison. Using a four-fold cross-validation strategy, we trained on 80% of the data and applied the best model to the remaining 20% for testing. The best performing model applied random forest and used phecode counts as input, with no dimensionality reduction. At a probability threshold of 0.5, this model correctly classified 392/452 (87%) of cases and 1,758/1,834 (96% of controls) while capturing 392/468 (84%) of cases and 1,758/1,818 (97%) of controls. Further, the model had an area under the receiver operator curve (AUROC) of 0.97 (Figure 1a) and an area under the precision recall curve (AUPR) of 0.92 (Figure 1b). Calibration was measured with a Brier score of 0.0460 after the application of isotonic regression (Figure 1c). Gini feature importance were largely correlated with the results from the pheWAS pointing to mostly developmental phenotypes (Supplementary Table 1).

**Figure 1.**
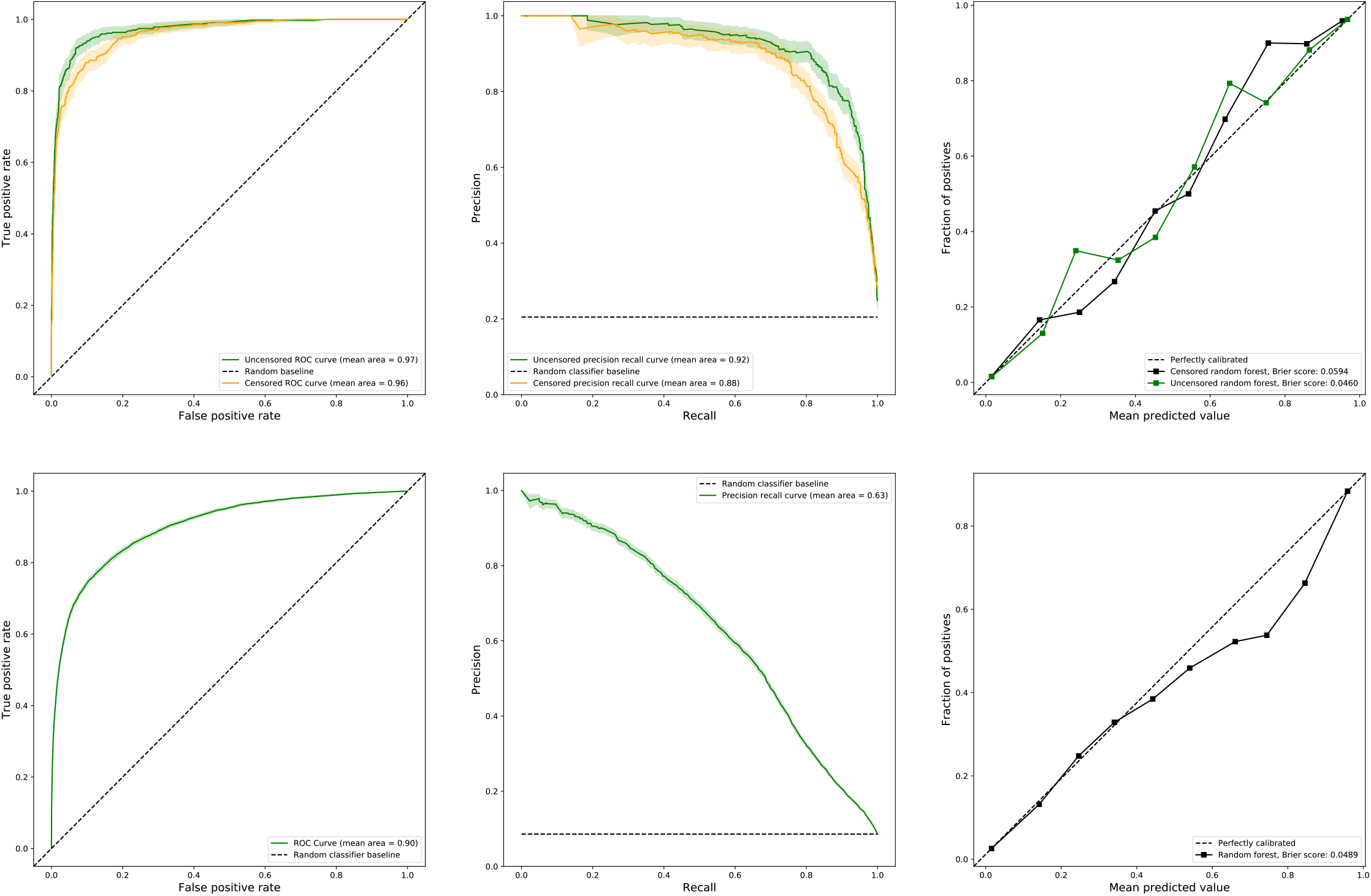
Performance metrics of genetic testing prediction model applied to held out test CMA dataset from both uncensored and censored versions (a-c) and applied to hospital population (d-f).

To assess whether model performance was biased by phecodes that occurred after the genetic test, we performed a secondary analysis in which we censored phecodes of CMA patients from the day their report was entered onwards. Despite a loss of phecode data (average time between first and last censored phecode: 686 days), the censored model still performed similarly to the uncensored model (AUROC = 0.96, AUPR = 0.88, Brier score = 0.0594, Figure 1a-c). We therefore use the uncensored model going forward. Finally, we assessed model disparity by building models using the same input data to predict self-reported race and sex. These models performed poorly compared to our model to predict genetic testing with much lower AUROCs (sex: 0.72 and race: 0.67) and AUPRs (sex: 0.62 and race: 0.22). However, they performed better than random and patients with high probabilities represented the distributions of race and sex among those in our training data which were disproportionately white and male (Table 1).

### Validation in a hospital reference population

CMAs are often the first line of genetic testing performed but do not account for all genetic testing in a hospital system. In order to validate our model on a broader set of patients receiving a genetic test, we applied it to a hospital sample that included 172,265 patients under 20 years of age (to match our training population) and having at least four years of data (Table 1). We defined cases as those having evidence of visiting a genetics clinic and controls as those with no mention or suspicion of genetic disease across their medical record (see Methods). In total, there were 10,074 cases and 107,263 controls. Applying the model in this population (Figure 1d-f) resulted in comparable classification performance (AUROC = 0.9) but lower average precision to the CMA test dataset which is at least partially driven by the much larger case imbalance (AUPR: 0.63).

### Genetic validation among CNV carriers

CNVs were generated from genotyping data on an independently ascertained subset of 6,445 patients from our hospital population described above (see Methods). We assessed the model’s performance in identifying patients with known or putative pathogenic variants in three ways. First, we identified 132 patients that carried a 10 Mb or greater duplication on chromosome 21. Based on diagnostic codes and explicit mentions in notes all of these patients had a clinical diagnosis of Down syndrome (DS), validating the CNV calls. Among these patients, the median probability was 0.92 (mean = 0.82) and 118 (89%) had probability greater than 0.5. The 15 patients with probabilities below 0.5 had four-fold fewer phecodes (mean: 174.4, mean unique: 24.8) compared to those with probabilities greater than 0.5 (mean: 698.1, mean unique: 65.1).

Second, patients were defined as having a CNV syndrome if they carried a deletion or duplication overlapping at least 50% of one of 23 highly penetrant, recurrent, pathogenic (Grade I) CNV syndromes from Decipher (22 deletions, 1 duplication)^22^. There were 46 patients, including 44 carrying deletions and 2 carrying duplications, that met this criterion (Figure 2a). The median probability in these patients was 97% (mean = 82%) with 40 (87%) having probability above 0.5 and 31 (67%) above 0.9. Nine syndromes were represented in this group with the most frequent including DiGeorge syndrome, Angelman/Prader-Willi syndrome, and Cri du Chat syndrome. Of the 6 patients with probabilities below 0.5, 2 had CNVs associated with neuropathies that typically present with symptoms later in life. Among our CMA sample, these patients received their reports when older than 10 years old on average compared to near birth for diseases like Down syndrome or DiGeorge syndrome. (Supplementary Fiigure 2). For one of the neuropathies, Hereditary Liability to Pressure Palsies (HNPP) we see a diverse presentation of symptoms corresponding to more variable predictions that may be a product of age (Figure 2b).

**Figure 2.**
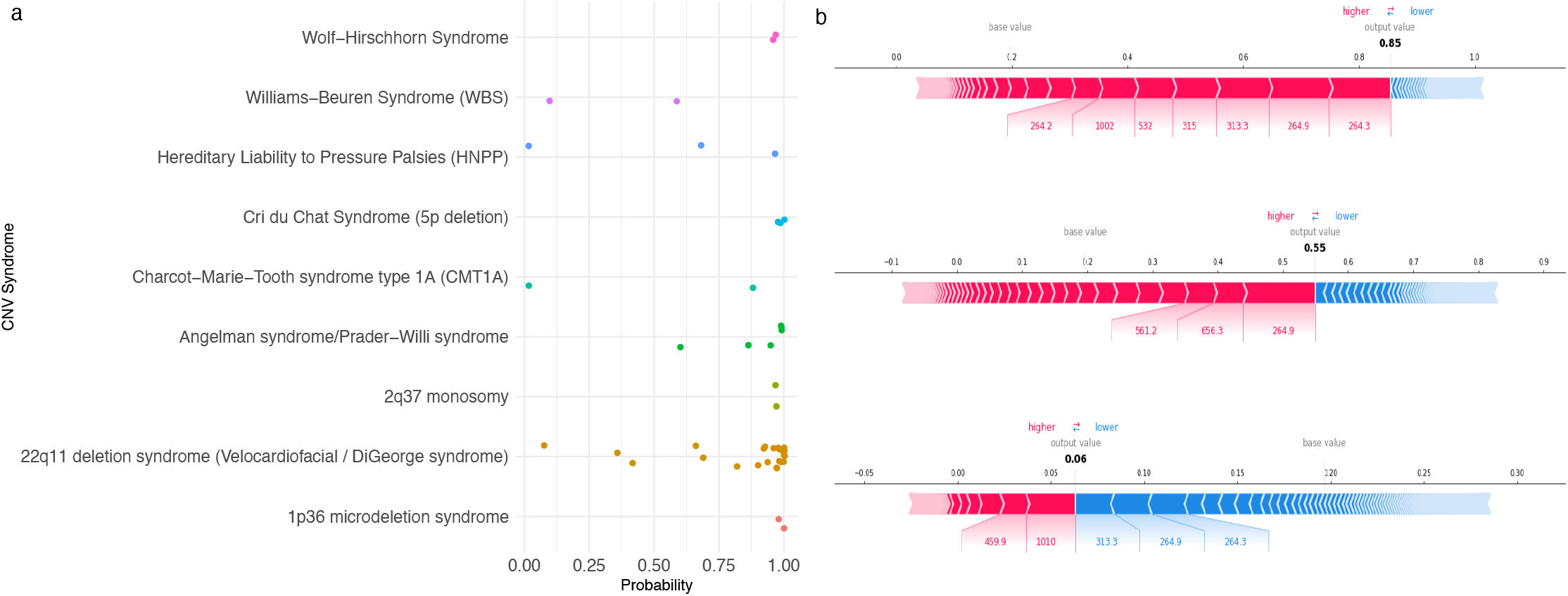
Probabilities for each of the 46 patients in the hospital sample with an overlapping pathogenic CNV syndrome stratified by specific disease (a). Tree Explainer plots^23^ for all 3 HNPP patients showing the phecodes that contribute to the posterior probabilities from the random forest model. Probabilities are before recalibration (i.e. decision scores), blocks represent a phecode, red implies contributes to increased probability, blue implies contributes to decreased probability with amount of contribution represented by the width.

Finally, we identified patients with at least one CNV overlapping 50% of a pathogenic CNV from ClinGen (n=7,773). This is a much larger set of curated pathogenic variants that we can use to quantify the proportion of patients with a possible genetic disease captured at different probability thresholds as well as how many patients appear to be undiagnosed. In total, 673 patients (10%) had at least one CNV overlapping at least one of these variants. The proportion of patients carrying a putative pathogenic variant increased to over 22% as the probability threshold increased (Figure 3). For comparison, 15.2% of the CMA patients had a reported abnormal gain or loss that overlapped 50% of a ClinGen pathogenic CNV. Further, 435 (64.6%) of these patients had no known interaction with the healthcare system for genetic reasons and 152 had probabilities greater than 0.5 marking a population captured by our model that will be highly enriched for genetic diseases but lack any current testing or intervention. Across the entire hospital population, there are thousands of patients with evidence of needing a genetic test but no record of seeing a genetics provider (n = 10,979 at probability > 0.5) or any mention of genetics issues in their notes (n = 2,238 at probability > 0.5).

**Figure 3.**
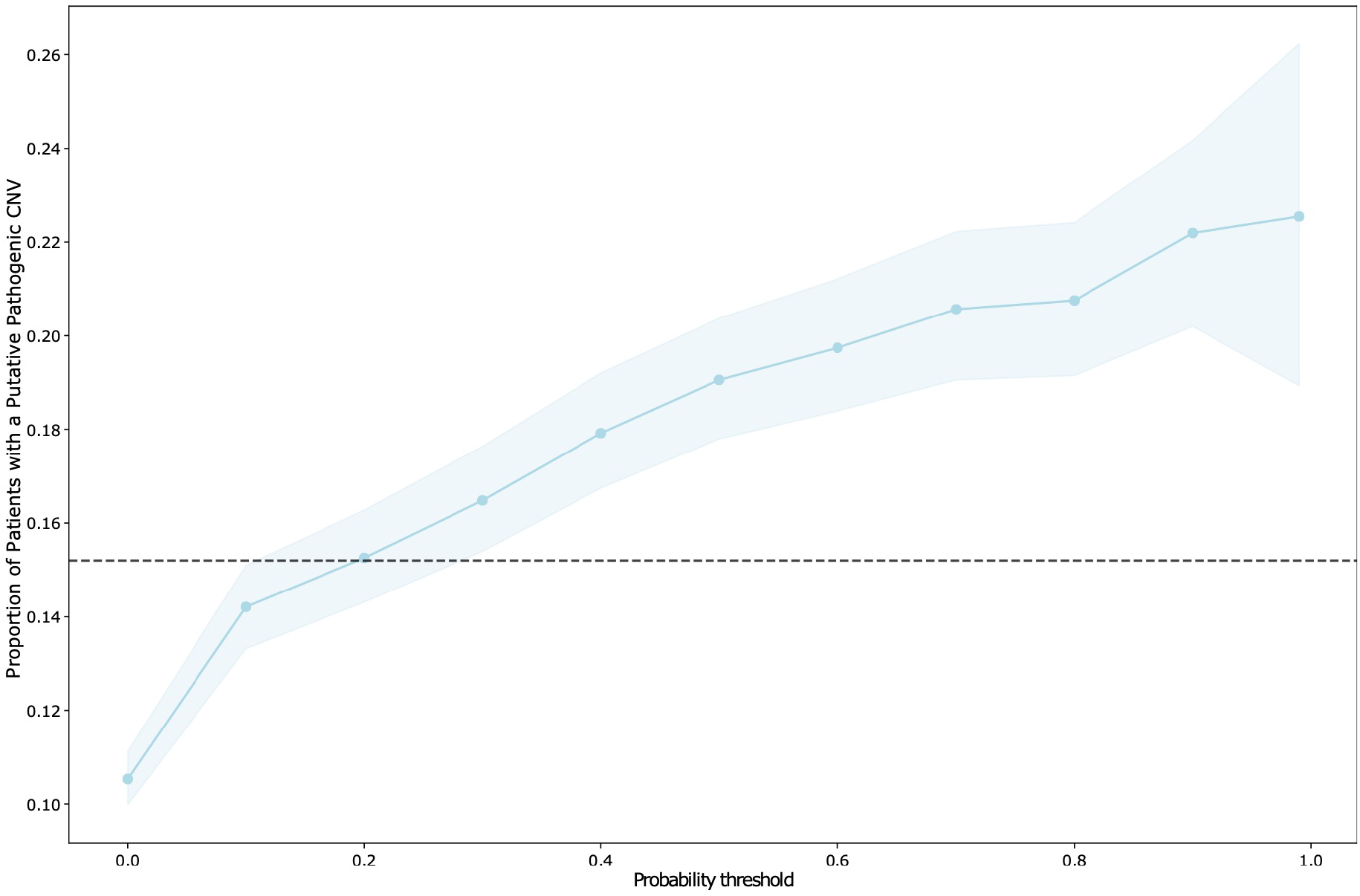
Proportion of patients with a CNV overlapping at least 50% a putative pathogenic CNV in ClinGen stratified by probability threshold. Dashed line represents proportion of reported abnormal gains or losses from CMA patients that overlap at least 50% of a ClinGen CNV. Colored band represents the bootstrapped 95% confidence interval.

### Model performance on patients with a diverse set of 16 genetic diseases

There are numerous genetic diseases which would not be included in our training dataset since a CMA would not be the appropriate genetic test. To assess our hypothesis more broadly, we tested our model’s ability to predict patients with a diverse set of 16 genetic diseases previously identified and validated in our sample^15^. These genetic diseases were selected for occurring frequently and for being well characterized for EHR based work. They ranged from syndromes based on large genomic alternations such as Down syndrome, DiGeorge syndrome, and fragile X syndrome of which some individuals existed in our training dataset to many other common genetic diseases such as cystic fibrosis, hemochromatosis, and sickle cell anemia which would not be present in our training dataset. In total, 1,843 patients in our hospital population had a chart validated diagnosis of at least one of these diseases. On average, our model identified the entire group of patients 4-8 times more frequently than expected based on the population rate of testing at different probability thresholds (Figure 4). For example, 1,051 patients had a probability greater than 0.5 corresponding to 57% of those with a diagnosis of one of these diseases whereas only 9% of the population would be tested at this threshold (6x increase in identification). Model performance was best on the syndromes caused by large genomic alterations capturing 76% of these patients at probability threshold of 0.5. However, regardless of genetic architecture and whether a disease was included in training, all of these disorders are captured better than population expectation with several including tuberous sclerosis, cystic fibrosis and Duchenne’s muscular dystrophy being particularly well captured at most thresholds (Figure 4).

**Figure 4.**
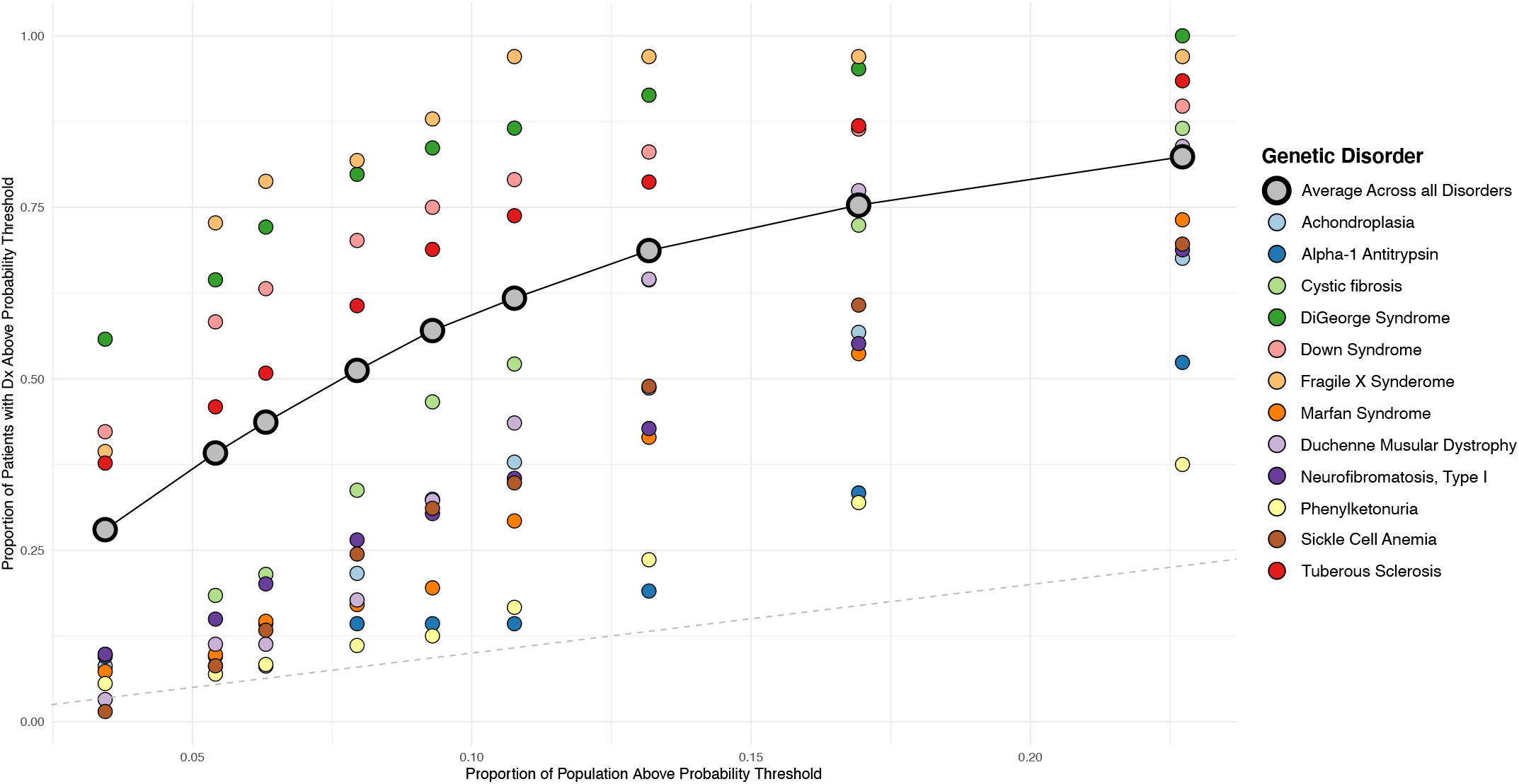
Proportion of patients diagnosed with one of 16 genetic diseases above a probability threshold (x-axis) compared to the proportion of patients that would be tested above the same probability threshold (y-axis). Large grey dots are values across all 16 genetic diseases. Among the 16 diseases, the 12 with more than 20 cases were plotted individually. The dashed line represents the identity line where the proportion of cases above threshold is equal to the proportion of the sample tested above that threshold. The 9 probability threshold increase from right to left and represent (>0.1, >0.2, >0.3, >0.4, >0.5, >0.6, >0.7, >0.8, >0.9).

## Discussion

Thousands of genetic diseases have been described based on presentation of a set of phenotypes seen across multiple individuals. While the specific profile of phenotypes may be unique, the overall pattern of multiple rare phenotypes that indicates a genetic disease is shared. Here, we show that this pattern can be predicted from phenotype data in the EHR, in essence, demonstrating the potential to automate and systematize clinical suspicion of a genetic disease that is the primary indication for getting a genetic test. We further validate the ability of this prediction model to identify patients who received a genetic test, not just a CMA, in a real-world population of hospital patients and those having genetic diseases based on clinical diagnosis or genetic evidence.

Genetic testing is crucial for diagnosis, prognosis and treatment or rare diseases. Yet, it is not consistently or equitably provided to those who need it and is often delayed by many years when it is offered. Our work here demonstrates the potential of using EHR data and machine learning to systematically identify patients that should receive a genetic test. Our results point to thousands of patients with phenotypes indicating the need for a genetic test but having no clinical suspicion in their medical record. A substantial number of these patients might finally receive a genetic diagnosis with the potential to alter their care. Further, this type of approach could lead to identification of new genetic diseases and improved phenotypic understanding of previously identified ones. Implementation of this type of model as an additional piece of information contributing to clinical suspicion could reduce time to testing, identify undiagnosed patients, and flag unnecessary tests, thereby improving care and reducing costs.

Using a set of putative pathogenic CNVs we were able to show that the proportion of patients who would have a pathogenic finding reached over 20% at higher probability thresholds. This proportion compares favorably to the 15.2% of our CMA patients that had an abnormal gain or loss variant overlapping the same set of CNVs. Importantly, our model identifies 10,979 patients with high probabilities (>0.5) and no recorded interaction with a genetics provider and 2,234 patients who have high probabilities (>0.5) yet lack any clinical suspicion of a genetic cause. These results indicate that implementation of such a model would provide at least as good a diagnostic yield as the current determination of genetic testing while more completely capturing those that could benefit from testing. While the model was trained on patients receiving a CMA, which is typically the first line test, we wanted to assess the model’s ability to identify patients with other genetic diseases for which a CMA would not be the appropriate test. Despite the specific nature of the training data, when validating the model among a set of 16 genetic diseases performance for many of the diseases that the model was not trained on was still high. This result points to the importance of our hypothesis, the consistency of that pattern of many rare phenotypes across many genetic disorders and the broader applicability.

An ongoing goal of this work is to directly improve prediction of patients with a genetic disease. In our training dataset, about 20.6% of those receiving a CMA reported an abnormal gain or loss. While this provides a subset which we could have trained on, there are two important limitations. The first is that all of these patients were ascertained based on the same clinical suspicion of having a genetic disease, and therefore needing a CMA. In fact, there are minimal phenotypic differences between those with an abnormal CMA and those without for that exact reason. Further, a CMA only has resolution to identify large genetic alterations, which are more likely to be of high effect but are less frequent than variants of smaller size that could also have large effect. In order to enable a model which can directly inform likelihood of carrying a genetic disease we will require higher resolution genetic data such as genome sequencing and a full clinical assessment of pathogenicity. This type of effort is ongoing and these data will be used to amend the training data in order to improve the model and move towards predicting genetic disease.

There are several limitations to note in this work. The current model is trained exclusively on young patients (< 20 years of age) most frequently having developmental issues with suspicion of carrying large chromosomal anomalies. There are many genetic diseases that would not receive this particular test and therefore would be excluded from our training data. While our model performs better than expected for a diverse set of 16 diseases, it performs better for diseases most similar to those it was trained on, particularly at the highest probabilities. We anticipate substantial improvements in performance and expansion to a larger population will be made when incorporating additional genetic data into the training of the model. It is important that any model built into healthcare not have explicit biases and that our algorithm is fair^24^. We tested whether the data going into our model could predict sex or race. While the prediction performance for these features was substantially worse than for our intended outcome of genetic testing it was not equivalent to a random model. This implies that although the model was unaware of race and sex, combinations of features still encoded this information, so it is not blind to these attributes. Our training data is skewed to higher proportions of males and of white individuals which is contributing to those populations having higher probabilities overall. Based on epidemiological data, it is expected that males will be at higher risk for the developmental disorders that are most commonly tested by CMA so this increased rate may be biological and appropriate. However, it is not clear that the increase in probabilities for white patients is appropriate and further work is needed to ensure any such model is not increasing disparities in healthcare before implementation. Finally, this approach requires longitudinal EHR data, and as seen in a subset of patients with Down syndrome when data is limited it could negatively affect performance. Additional work is required to assign confidence to these predictions based on the amount and specific phenotype data available for a given patient. Importantly, the current model only uses structured diagnostic codes making it more amenable for use within many other systems.

In conclusion, we present an approach that leverages EHR data and machine learning to predict which patients should receive a genetic test based on the hypothesis that a unique constellation of rare phenotypes is a hallmark feature of genetic disease. We show that this model can accurately predict patients needing a genetic test across multiple datasets, using differing definitions of genetic tests, among patients carrying pathogenic CNVs and across numerous genetic diseases. There exists the potential for a model of this type to improve the healthcare of those with genetic diseases by speeding up diagnosis and reducing healthcare burden and costs.

## Data Availability

Code and scripts are available at https://github.com/RuderferLab/chromosomalMicroarray

## Author contributions

DMR and TJM designed and conceived the study. LB and TJM extracted data from the EHR for training and validation. LH generated the CNV data. TJM, DMR and JM designed and implemented the prediction model. TJM and DMR performed analyses. DMR, TJM, LB and NJC provided interpretation of results. TJM and DMR drafted the manuscript. All authors have read, provided feedback and approve submission.

## Acknowledgements

This work was supported by R01MH111776 (DMR), R01MH113362 (NJC, DMR), R01LM010685 (LB), and U01HG009068 (NJC). This study makes use of data generated by the DECIPHER community. A full list of centres who contributed to the generation of the data is available from https://decipher.sanger.ac.uk and via email from. Funding for the project was provided by Wellcome. The dataset(s) used for the analyses described were obtained from Vanderbilt University Medical Center’s BioVU which is supported by numerous sources: institutional funding, private agencies, and federal grants. These include the NIH funded Shared Instrumentation Grant S10RR025141; and CTSA grants UL1TR002243, UL1TR000445, and UL1RR024975. Genomic data are also supported by investigator-led projects that include U01HG004798, R01NS032830, RC2GM092618, P50GM115305, U01HG006378, U19HL065962, R01HD074711; and additional funding sources listed at https://victr.vanderbilt.edu/pub/biovu/.

